# A Genetic Profile of Refractory Individuals with Major Depressive Disorder and Their Responsiveness to Transcranial Magnetic Stimulation

**DOI:** 10.1101/2020.04.13.20063404

**Authors:** Nathália G. Souza-Silva, Eduardo S. Nicolau, Kate Hoy, Ziarih Hawi, Mark A. Bellgrove, Débora M. Miranda, Marco A. Romano-Silva, Paul B. Fitzgerald

## Abstract

**Background:** Major depressive disorder (MDD) is a debilitating illness characterized by the persistence of negative thoughts and emotions. Although antidepressant medications are effective, less than half of patients achieve complete remission despite multiple treatment trials. Repetitive transcranial magnetic stimulation (rTMS) has proven effective in the treatment of depression, especially for patients resistant to antidepressant medications. Remission rates when using rTMS for treatment-resistant depression (TRD) patients are between 30% and 40%. The responsiveness to pharmacotherapy and rTMS therapy may be influenced by genetic factors.

**Objective:** Here we aim to characterize the genetic profile of refractory individuals with MDD and their rTMS responsiveness.

**Methods:** We used an extreme-phenotype design (rTMS responders vs. non-responders) and conducted a genome wide association study on 48 participants and 593,260 SNPs.

**Results:** We identified 53 significant SNP associations. Gene-set enrichment analysis showed that significantly associated genes loaded onto synaptic plasticity regulation pathways. Among the genes found differentially expressed in rTMS responders compared to non-responders were *APP, GRID2* and *SPPL2A* genes.

**Conclusions:** Based on these findings, we suggest that the identified genes may influence of rTMS responsiveness. Furthermore, the rTMS responsiveness may be associated with several pathways and not just to the influence of a single gene. To the best of our knowledge, this is the first report on the genetic profile of rTMS response using a GWAS approach. Nevertheless, further studies are necessary to enlight the molecular mechanism by which these genes affect response to rTMS treatment.

## Introduction

Major depressive disorder (MDD) is characterized by the persistence of negative thoughts and emotions that disturb mood, cognition, motivation and behavior [1]. According to the World Health Organization (WHO) [2] depression is the main reason of disability worldwide, affecting around 300 million people. Depression can occur at any stage of life, from childhood until old age, with a twofold higher incidence in women compared to men [3]. Several treatment options are available for depression, such as pharmacological and non-pharmacological therapy, psychotherapy and brain stimulation techniques. However, less than half of patients with MDD achieve complete remission after a first line treatment with antidepressants [4].

Treatment-resistant depression (TRD) refers to the occurrence of an inadequate response after antidepressant treatment among patients that suffer from unipolar depressive disorder [5]. The most traditional concept of TRD is based on the non-responsiveness to at least two protocols of antidepressant therapy [6]. Among the patients who receive adequate treatment for MDD, only 30% respond to treatment and achieve remission. Of the remaining 70%, approximately 20% of the patients present an improvement in depressive symptoms, although not achieving remission, while 50% do not present any kind of response [7]. Due to the low efficacy of antidepressants after two attempts of treatment without success, new alternative therapies have been developed. The use of neurostimulation strategies are potential candidates acting as alternative or complementary therapies for neuropsychiatric disorders.

Repetitive transcranial magnetic stimulation (rTMS) has been demonstrated to be an effective depression treatment [8]. In rTMS, electromagnetic induction is used to induce focal currents in superficial brain regions and modulate cortical function [9]. Previous studies have demonstrated that low-frequency stimulation of rTMS (≤ 1 Hz) leads to cortical activity inhibition, whereas high-frequency stimulation (≥ 5 Hz) increases cortical excitability [10]. Although rTMS is an effective treatment for many patients with TRD, it is not always effective with remission rates ranging from 30% to 40% and response rate between 45% and 60% [11]. The factors contributing to rTMS responsiveness remain unclear. Although one potential source of inter-individual variability in responsiveness to rTMS could be genetics, few studies have sought to identify a genetic basis of rTMS response [12].

The genetic basis of depression is well established through twin and family studies with heritability estimates ranging from 30% to 40%, and SNP-based heritability estimates ranging from 9% to 29% [13,14]. Risk of MDD is highly polygenic and involves many genes with small effects. This coupled with the clinical heterogeneity of MDD requires very high numbers of patients to find significant associations [15]. Genome-wide association study (GWAS) is a powerful tool for investigating the genetic risk factors of complex human disease, providing information about variants that may be associated with a trait [16,17]. GWAS has been used to map genetic loci, associated with MDD [18,19]. A recent GWAS conducted by Wray et al (16,823 MDD cases and 25,632 controls) identified 44 risk variants and significant loci associated to MDD [14]. Only hypothesis driven SNP genotyping approaches have been so far applied in studies with rTMS response, mainly focusing on BDNF [20–23] or serotonergic genes such as 5-HTTLPR [24].

In face of such knowledge, we hypothesized that inter-individual differences in genetics may influence the responsiveness of rTMS in patients with treatment resistant MDD. To explore this hypothesis we used an extreme-phenotype design in which we compared allelic variation genome-wide between rigorously defined rTMS responders and non-responders.

## Materials and Methods

### Participants

The study was approved by the Human Research and Ethics Committee of the Alfred Hospital. All patients had a DSM IV diagnosis of major depressive disorder applied by an experienced psychiatrist and confirmed by the Mini-International Neuropsychiatric Interview (M.I.N.I) [25]. Patients who received rTMS treatment whilst participating in one of two clinical trials [26,27] were recruited for genetic analysis. All patients received high frequency left sided 10Hz rTMS, either in a standard daily format or in a more accelerated treatment protocol (Table 01). Patients were asked to donate saliva for DNA samples. The DNA extraction was conducted using a standard protocol as recommended by the supplier Oragene® (Kit Oragene-DNA | OG-600 prepIT-L2P). The resulting purified gDNA is ideal for microarray analysis.

**Table 01.** Characteristics of participants. Abbreviations: SSRI, Selective Serotonin Reuptake Inhibitor; MAOI, Monoamine Oxidase Inhibitor; SNRI, Serotonin-norepinephrine Reuptake Inhibitor; TCA, Tricyclic Antidepressant; RIMA, Reversible Inhibitor of Monoamine Oxidase-A; NaSSA, Noradrenergic and Specific Serotonergic Antidepressant.

| **Age - Mean (SD)** |  |
| --- | --- |
| Age (years) | 47 (13,26) |
| Range (years) | 19-74 |
| **Gender (%)** |  |
| Male (#) | 21 (43,75) |
| Female (#) | 27 (56,25) |
| **Occupational status (%)** |  |
| Employed (#) | 15 (31,25) |
| Unemployed (#) | 21 (43,75) |
| Part-time (#) | 6 (12,5) |
| Retired (#) | 3 (6,25) |
| N/A (#) | 3 (6,25) |
| **Age onset (%)** |  |
| Childhood (#) | 5 (10,42) |
| Adolescence (#) | 15 (31,25) |
| Early adulthood (#) | 16 (33,33) |
| Mid adulthood (#) | 12 (25) |
| **Use of antidepressant (%)** |  |
| SSRI | 8 (16,66) |
| MAOI | 3 (6,25) |
| SNRI | 14 (29,16) |
| TCA | 3 (6,25) |
| RIMA | 1 (2,08) |
| NaSSA | 1 (2,08) |
| Combination | 7 (14,58) |
| None | 9 (18,75) |
| N/A (#) | 2 (4,16) |

A total of 99 (100%) individuals consented and provided samples for analysis. Clinical outcomes (response or non-response) were determined based on scores on the Montgomery Asberg Depression Rating Scale (MADRS). We compared MADRS scores from baseline to the end of acute treatment. We included individuals for analysis who were either clear responders to rTMS (a greater than 60% reduction on the MADRS scale – N = 29 (29.29%) or clear non-responders (below a 10% reduction on the MADRS scale – N = 19 (19.19%)). The remaining 51 (51.51%) subjects who demonstrated a reduction of between 11-59% were excluded from the analysis. These criteria were applied considering that extreme scoring patients (<10 >60) may present more representative genetic results to the allocated groups. Recently association studies have been sampling the extremes as a strategy for achieving good statistical power under sample size limitations. This strategy is based on the assumption that extreme phenotype sampling may increase power to detect causal variants [28–30].

### TMS Treatment

Treatment followed the same conditions described by Fitzgerald *et al* [26,27]. In one study all patients received a 3 week course of 10 Hz stimulation applied to the left DLPFC with an extension of this course up to 6 weeks in total or switching to low frequency right sided rTMS or bilateral rTMS if not meeting partial response criteria at 3 weeks [26]. Patients in the second study received one of two treatment conditions – accelerated rTMS and standard rTMS. In the accelerated treatment, in week 1, patients were provided 3 sessions per day over 3 days. In week 2, patients were provided 3 sessions per day over 2 days and in week 3, 3 sessions in a single day were provided. In a standard treatment, 20 daily sessions provided 5 days per week over 4 weeks [27]. Both treatments provided trains of 10 Hz rTMS to the left dorsolateral prefrontal cortex (DLPFC).

### Genotyping

Genotyping was performed using the Infinium PsychArray-24 BeadChip (Illumina, Inc., San Diego, CA, USA) and automated workflow according to the manufacturer’s instructions. Raw data were analyzed using PLINK 1.9 [31].

### Data quality control

Due to large number of marker loci tested in GWAS a rate of error or bias can be harmful. Therefore, to remove false-positive or false-negative associations, steps of quality control was performed to remove individuals or markers with high error rates. Data quality control was performed using PLINK 1.9 [31]. SNP inclusion required: call rate (GENO) > 90%, maximum individual missingness rate (MIND) > 10%, minor allele frequency (MAF) < 5% and Hardy-Weinberg Equilibrium (HWE) p-value > 10^-6^.

### Association analysis

For the analysis of association between the phenotype and the response to rTMS therapy we used the resources available in PLINK 1.9 [31]. We performed standard association analysis to compare allele frequency in both groups (responders and non-responders) with a 95% confidence interval through the following commands (--assoc), (--ci 0.95) (--adjust).

### Pathways Analysis

The protein-protein interaction (PPI) network analysis of the identified hub genes was constructed using the Search Tool for the Retrieval of Interacting Genes (STRING) database (database of known and predicted protein-protein interactions) [32]. The resulting PPI network was then visualized using Cytoscape [33] software (software platform for visualizing molecular interaction networks and biological pathways that integrating these networks with annotations, gene expression profiles) (ClueGO plug-in) for the functional enrichment analysis.

## Results

Since the observed clinical responses between both trials were similar, data analysis and results were presented in conjunction (Table 02). Quality control analysis on the raw genotypic data was conducted in 48 individuals and 593,260 SNPs. After application of data quality control criteria, 958 variants were removed due to missing genotype data, 310,522 SNPs were removed due to minor allele threshold and 4 people were removed due to missing genotype data. This left 281,780 SNPs and 44 subjects for the association study. In order to estimate the effective number of significant SNPs, we submitted the results to the False Discovery Rate (FDR) correction considering sample and SNP size per chromosome [34,35]. A new *p* value was then determined for each chromosome (Table 03). GWAS analysis using PLINK 1.9, revealed 53 significantly SNP associations, 11 of which were related to treatment response and 42 associated with non-responsiveness to treatment. Of the 53 associated SNPs, 25 were mapped to non-coding genomic regions. The remaining 28 SNPs mapped to protein coding genes; 9 SNPs mapped to described pathways (Table 04).

**Table 02.**
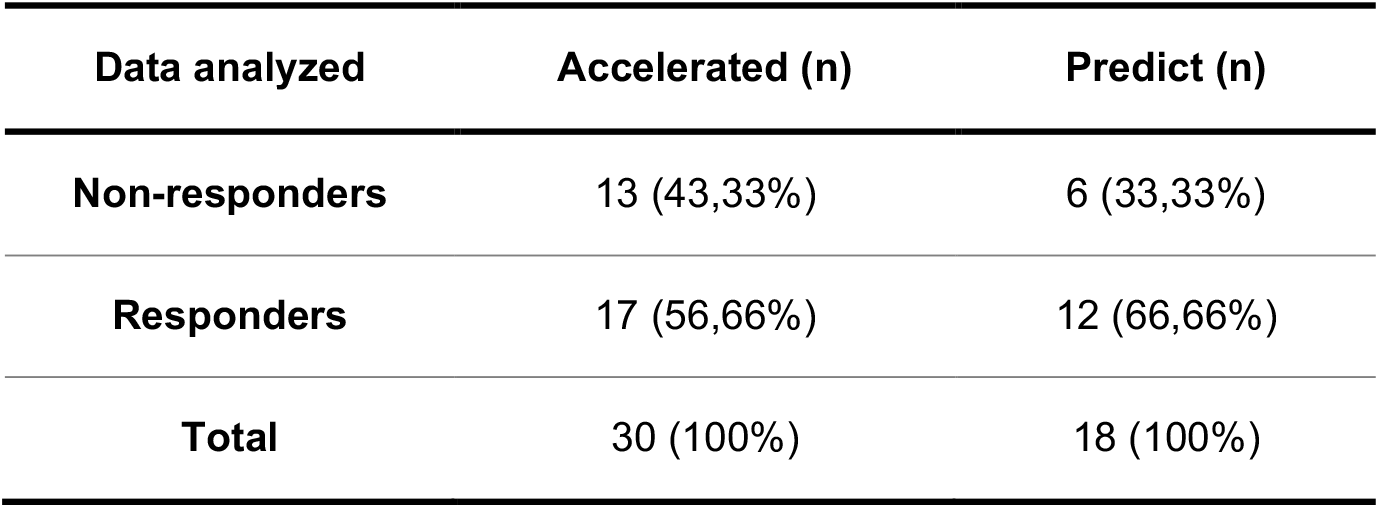
Response rates after treatment with rTMS. Analysis conducted in GraphPad Prism with Chi-square Test and Fisher’s exact test to show the difference between groups after treatment with rTMS (*p* 0.5544).

**Table 03.** New p value per chromosome after FDR correction.

| Chr | Total SNP | New *p*-value (FDR) |
| --- | --- | --- |
| 1 | 19934 | 0.000106 |
| 2 | 20574 | 0.000090 |
| 3 | 17491 | 0.0000936 |
| 4 | 16104 | 0.0001141 |
| 5 | 15623 | 0.0001225 |
| 6 | 17610 | 0.0001126 |
| 7 | 14047 | 0.0001552 |
| 9 | 11283 | 0.0001893 |
| 10 | 12745 | 0.0001457 |
| 11 | 12742 | 0.0001628 |
| 12 | 12093 | 0.0001572 |
| 13 | 9490 | 0.000218 |
| 14 | 7895 | 0.0002183 |
| 15 | 7537 | 0.0002380 |
| 17 | 7233 | 0.0002747 |
| 18 | 7408 | 0.0002840 |
| 19 | 5896 | 0.0003472 |
| 21 | 3703 | 0.0004098 |
| 22 | 3687 | 0.0004761 |
| X | 6442 | 0.0001432 |

**Table 04.** SNPs founded. Description of significant SNPs (p <0.05). A1, lower frequency allele. A2, highest frequency allele. MAF, minor allele frequency. SNV, single nucleotide variant. OR (0dds ratio) > 1 related to treatment response and OR < 1 associated to non-responsiveness treatment.

| Chr | SNP | Position | Gene | Chi square | Odds Ratio | P value | A1 | A2 | MAF |
| --- | --- | --- | --- | --- | --- | --- | --- | --- | --- |
| 19 | rs960995 | 57039169 | *ZNF471* | 13.49 | 0.18 | 0.0002397 | G | A | 0.4886 |
| 7 | rs17164813 | 11616500 | *THSD7A* | 16.54 | 0.1373 | 0.000047516 | A | C | 0.3068 |
| 15 | rs8035452 | 51040798 | *SPPL2A* | 14.53 | 7.25 | 0.0001376 | G | A | 0.3977 |
| 18 | rs595562 | 18449508 | snv variation<br>near genes<br>*LINC01541* and<br>*LOC107985179* | 15.50 | 0.1603 | 0.000082719 | G | A | 0.3714 |
| 1 | rs4648426 | 3773089 | snv variation<br>near genes<br>*DFFB* and<br>*CEP104* | 14.43 | 0.1489 | 0.0001455 | G | A | 0.2727 |
| 3 | rs12487861 | 160535721 | *PPM1L* | 18.38 | 16 | 0.000018101 | A | C | 0.3295 |
| 11 | rs198475 | 61526071 | *MYRF* | 14.43 | 0.1489 | 0.0001455 | A | G | 0.2727 |
| 1 | rs560681 | 160786670 | *LY9* | 14.98 | 0.09502 | 0.0001088 | G | A | 0.1818 |
| 1 | rs11265485 | 160764759 | *LY9* | 16.98 | 0.08403 | 0.000037682 | G | A | 0.1932 |
| 9 | rs1934115 | 23103266 | *LOC107987055* | 14.49 | 0.04528 | 0.000141 | C | A | 0.125 |
| 18 | rs872994 | 73171838 | *LOC107985177* | 13.919 | 0.1769 | 0.0001909 | A | C | 0.375 |
| 22 | rs5995416 | 37719004 | *LOC105373024* | 12.643 | 5.61 | 0.0003769 | A | G | 0.4432 |
| 19 | rs2189698 | 57014071 | *LOC105372471* | 13.52 | 0.1839 | 0.0002364 | C | A | 0.4318 |
| 18 | rs4243296 | 73219777 | *LOC105372202* | 13.92 | 0.1769 | 0.0002 | G | A | 0.375 |
| 6 | rs6899975 | 138275769 | *LINC02528* | 14.38 | 0.1729 | 0.0001494 | A | G | 0.3977 |
| 17 | rs1014129 | 49517224 | *LINC02073* | 17.10 | 0.1407 | 0.000035449 | A | G | 0.3523 |
| 13 | rs626904 | 39984946 | *LHFPL6* | 14.90 | 0.1674 | 0.0001131 | A | G | 0.4205 |
| 2 | rs17673232 | 144860827 | *GTDC1* | 19.07 | 0.07451 | 0.000012614 | A | G | 0.2045 |
| 4 | rs11942069 | 94494455 | *GRID2* | 14.43 | 0.1489 | 0.0001455 | A | G | 0.2727 |
| 14 | rs447347 | 89992265 | *FOXN3* | 14.79 | 0.1654 | 0.0001205 | A | G | 0.4773 |
| 13 | rs2271926 | 39979675 | *EXOSC7* | 14.90 | 0.1674 | 0.0001131 | G | A | 0.4205 |
| 19 | rs4646515 | 15658569 | *CYP4F3* | 9.122 | 0.2503 | 0.002525 | G | C | 0.3636 |
| X | rs2273081 | 4594630 | *COL9A3* | 12.17 | 0.1571 | 0.0004864 | C | A | 0.3429 |
| 22 | rs229526 | 47236880 | *C1QTNF6* | 16.40 | 0.1324 | 0.00005141 | G | A | 0.3409 |
| X | rs2980075 | 152794075 | *ATP2B3* | 11.62 | 0.1282 | 0.0006537 | C | A | 0.2286 |
| 21 | rs373521 | 27257660 | *APP* | 12.5 | 0.1963 | 0.0004067 | A | C | 0.3864 |
| 6 | rs1283468 | 70038147 | *ADGRB3* | 15.56 | 0.07051 | 0.00007978 | A | G | 0.1591 |
| 5 | rs11956034 | 178754468 | *ADAMTS2* | 14.98 | 0.09502 | 0.0001088 | A | G | 0.1818 |
| 3 | rs501118 | 95116949 | - | 15.66 | 8.7 | 0.000075922 | A | G | 0.375 |
| 21 | rs9981074 | 33165958 | - | 13.54 | 6.29 | 0.0002341 | A | G | 0.4205 |
| X | rs17317597 | 116660237 | - | 8.202 | 12.55 | 0.004185 | G | A | 0.2143 |
| 18 | rs4347699 | 51183679 | - | 15.74 | 10.33 | 0.000072533 | A | G | 0.3409 |
| X | rs12559502 | 128048545 | - | 6.893 | 0.2462 | 0.008651 | G | A | 0.3 |
| X | rs17333434 | 27133302 | - | 8.072 | 0.2027 | 0.004495 | G | A | 0.2571 |
| 21 | rs2829964 | 27242396 | - | 12.823 | 0.1914 | 0.0003425 | A | G | 0.4659 |
| 21 | rs2142419 | 19928069 | - | 12.908 | 0.1778 | 0.0003272 | A | G | 0.3068 |
| 18 | rs8082822 | 73209334 | - | 13.92 | 0.1769 | 0.0001909 | A | G | 0.375 |
| X | rs6640653 | 4579981 | - | 9.714 | 0.1645 | 0.001829 | A | C | 0.2429 |
| 10 | rs2068888 | 94839642 | - | 15.50 | 0.1603 | 0.0000827 | G | A | 0.4432 |
| X | rs1343974 | 4594630 | - | 12.17 | 0.1571 | 0.0004864 | C | A | 0.3429 |
| 13 | rs9548721 | 39846266 | - | 14.569 | 0.1546 | 0.0001351 | G | A | 0.2955 |
| 12 | rs7135989 | 48655268 | - | 16.40 | 0.1324 | 0.000005141 | A | G | 0.2841 |
| X | rs5916687 | 4596138 | - | 11.62 | 0.1282 | 0.0006537 | A | G | 0.2286 |
| 21 | rs2829950 | 27223152 | - | 19.23 | 0.124 | 0.000011565 | C | A | 0.3636 |
| X | rs5980684 | 69300000 | - | 13.57 | 0.1222 | 0.0002298 | A | G | 0.2714 |
| 6 | rs1074349 | 22838984 | - | 18.63 | 0.1217 | 0.000015902 | G | A | 0.3182 |
| 22 | rs134913 | 27413509 | - | 18.41 | 0.1111 | 0.000017857 | G | A | 0.2727 |
| X | rs12390729 | 4597922 | - | 15.68 | 0.1053 | 0.00000055659 | A | G | 0.2857 |
| 13 | rs944868 | 39843411 | - | 18.47 | 0.102 | 0.000017268 | C | A | 0.25 |
| 10 | rs10787147 | 111079538 | - | 14.6 | 9.595 | 0.0001327 | G | A | 0.3295 |
| 14 | rs2094718 | 99434288 | - | 13.5 | 8.908 | 0.000238 | C | A | 0.3182 |
| X | rs1144863 | 144582143 | - | 12.83 | 7.986 | 0.0003411 | A | G | 0.4143 |
| X | rs5915786 | 4642016 | - | 9.775 | 5.353 | 0.001769 | G | A | 0.4571 |

Protein-protein interaction network (PPI) analysis performed through the STRING database, presented no pathway association between the identified genes (Table 05). In an attempt to explore an interaction network analysis between the selected genes, we included common pharmacological target genes for depression treatment in the analysis. Among the pharmacological target genes were *BDNF, COMT, SLC6A1*, however, no significant protein-protein interaction network was identified (Figure 01).

**Table 05.** Description of significant genes. Source: [69]

| Gene | Brain Expression | Function |
| --- | --- | --- |
| *SPPL2A* | + | Catalyzes the intramembrane cleavage of a several proteins and may play a role in the regulation of innate and adaptive immunity. |
| *APP* | + | Performs physiological functions on the surface of neurons relevant to neurite growth, neuronal adhesion and axogenesis. |
| *EXOSC7* | + | Presents exoribonuclease activity and participates in a multitude of cellular RNA processing and degradation events. |
| *GRID2* | + | Plays a role in synapse organization between parallel fibers and Purkinje cells. |
| *ADGRB3* | + | Plays a role in the regulation of synaptogenesis and dendritic spine formation. |
| *COL9A3* | + | Possesses the function of structural component of hyaline cartilage. |
| *LY9* | - | Modulates the activation and differentiation of a wide variety of immune cells and are involved in the regulation and interconnection of both innate and adaptive immune response. |
| *FOXN3* | + | Acts as a transcriptional repressor and may be involved in DNA damage-inducible cell cycle arrests (checkpoints). |

**Figure 01.**
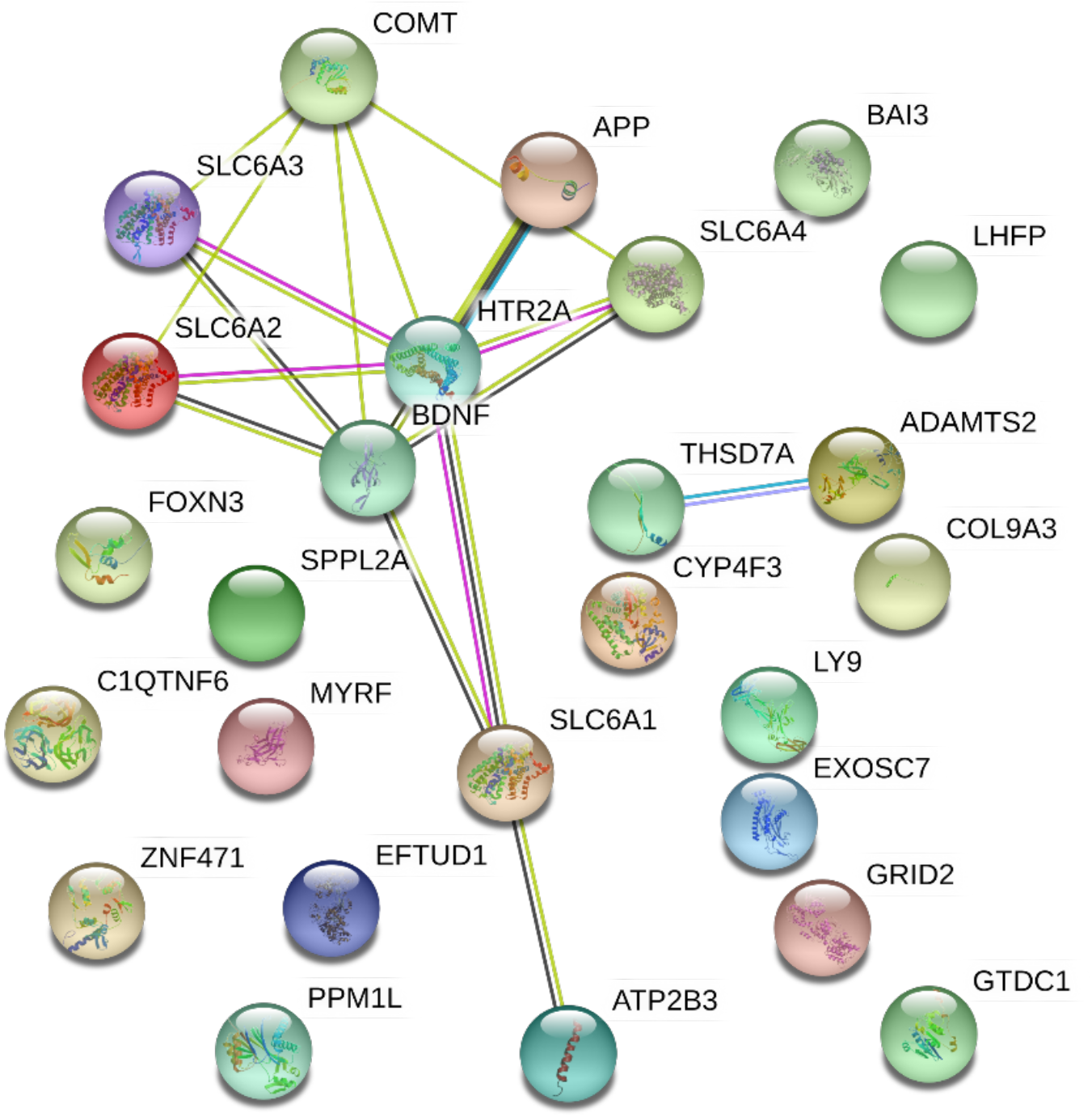
Protein pathway network from STRING. Genes of positive SNPs with pharmacological targets. Colors of edges: black (co-expression), green (textmining), pink (experimentally determined) and blue (from curated databases). Colors of nodes: colored nodes – query proteins and first shell of interactors, white nodes – second shell of interactors. Node content: empty nodes – proteins of unknown 3D structure, filled nodes – some 3D structure is known or predicted.

Genes and pathway interaction networks were obtained after enrichment analysis using ClueGo, a Cytoscape plug-in. The pathways involved were: synaptic plasticity regulation pathway, containing - *APP* (precursor beta amyloid protein), *SPPL2A* (signaling GPCR - transmembrane proteins), *GRID2* (glutamatergic ionotropic receptor), *ADGRB3* brain-specific angiogenesis inhibitor), *COL9A3* (structural constituent of the extracellular matrix) genes (Figure 02).

**Figure 02.**
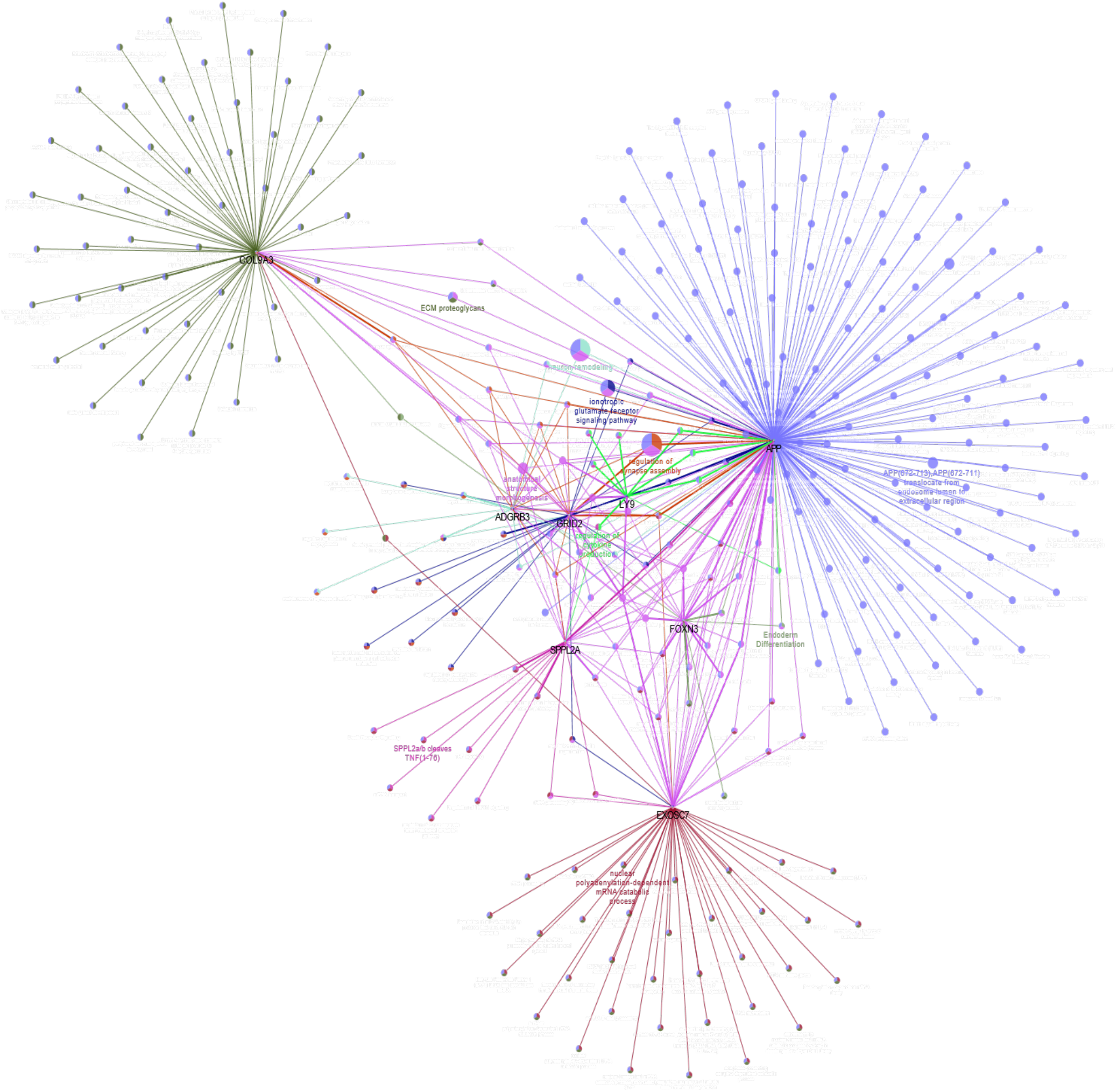
Regulation of synaptic plasticity pathway. In this way, the genes that are in bold and their hexagon-shaped nodules are the genes found after genome analysis through PLINK and Haploview. Ball-shaped nodules represent the pathways to which these genes participate. The interaction between the genes found results in the regulation pathway of synaptic plasticity. The genes involved in this pathway are: *APP* (amyloid beta protein precursor), *SPPL2A* (GPCR signaling - transmembrane proteins), *EXOSC7* (exosome component), *FOXN3* (forkhead/winged helix transcription factor family), *GRID2* (glutamatergic ionotropic receptor), *LY9* (Self-ligand receptor of the signaling lymphocytic activation molecule (SLAM) family), *ADGRB3* (brain-specific angiogenesis inhibitor), *COL9A3* (extracellular matrix structural constituent).

## Discussion

The results of this study reveal a number of SNPs that may be associated with the response to rTMS in treatment-resistant patients with major depressive disorder. Among the findings are genetic polymorphisms that have already been mapped to genes associated with the innate and adaptive autoimmune response, genes involved in the pathophysiology of Alzheimer’s disease (AD), regulation of synaptogenesis and dendritic spine formation [36–38]. Although these genes point to a contribution to the overall phenotype, it is important to elucidate the effects of each gene on disease development. It is worth mentioning that the sum of multiple genetic and environmental factors leads to different clinical presentations and therapeutic responses in each patient.

In our study, the repetitive TMS (rTMS) protocol to the left DLPFC was performed in predict and accelerated modalities. It was possible to observe that the treatment model adopted did not interfere in the individuals’ responsiveness to rTMS (Table 02). Therefore, for the genetic analysis patients from both protocols (predict and accelerated) were grouped.

Although we were not able to characterize a complete protein-protein interaction network with the SNP carrying genes, we were able to further describe their individual functionality and possible association to our disease and treatment in focus.

In the synaptic plasticity regulation pathway the significant genes found were related to the signaling of transmembrane proteins (*SPPL2A*), precursor of the beta amyloid protein (*APP*), exosomal component (*EXOSC7*), glutamatergic ionotropic receptor (*GRID2*), brain-specific angiogenesis inhibitor (*ADGRB3*), structural constituent of the extracellular matrix (*COL9A3*), lymphocyte antigen 9 (*LY9*) and forkhead box N3 (*FOXN3*) (Table 05).

### Positive Response Associated Genetic Variants

Signal peptide peptidase-like 2a (*SPPL2A*) is a gene that encodes an aspartic intramembrane protease that plays an important role in the development and function of antigen presenting cells such as B-lymphocytes and dendritic cells. Regulated intramembrane proteolysis (RIP) is a process that controls communication between cells and the extracellular environment mediated by a family of proteases, the intramembrane cleaving proteases (I-CLiPs) [39]. The founding members of this I-CliPs family are the presenilins (PS1 and PS2), the catalytically active subunit of the γ-secretase complex [40]. *SPPL2A* has been described as an enzyme related to presenilins [39]. The prominent class of γ-secretase is an aspartyl I-CLiPs involved in the generation of the beta-amyloid (Aβ) peptide from the amyloid precursor protein (*APP*). In AD patients it is possible to find the presence of amyloid plaques in neural tissue, and it is believed that the accumulation of these polypeptides is involved in the development of the disease [41]. Previous studies have shown that depression is one of the most frequent comorbid psychiatric disorders in Alzheimer’s disease and up to 50% of patients with AD will suffer from depression at some stage during the progression of dementia [42]. A study of Zhu *et al* [43] showed that similar environmental risk factors have been implicated in different neuropsychiatric diseases (including major depressive disorder and Alzheimer’s disease), indicating the existence of common epigenetic mechanisms underlying the pathogenesis shared by different illnesses.

### Negative Response Associated Genetic Variants

*EXOSC7* is a gene encoding RNA exosome – exosome component 7. The RNA exosome is a ribonuclease complex composed of both structural and catalytic subunits that participate in the processing of stable RNA species [44]. Mutational changes in genes encoding RNA exosome subunits may trigger inherited tissue-specific diseases [45]. A study conducted by Di Donato *et al* [46] showed that mutations in *EXOSC2* have been linked to a novel syndrome characterized by early onset retinitis pigmentosa, progressive sensorineural hearing loss, hypothyroidism, premature aging and mild intellectual disability. Other studies reveal that mutations in *EXOSC3* have been linked to pontocerebellar hypoplasia type 1 (PCH1b), an autosomal-recessive, neurodegenerative disease characterized by significant atrophy of the pons and cerebellum, Purkinje cell abnormalities, and degeneration of spinal motor neurons [47].

Glutamate ionotropic receptor delta type subunit 2 (*GRID2*) is a gene member of the family of ionotropic glutamate receptors which are the predominant excitatory neurotransmitter receptors in the mammalian brain. Single nucleotide polymorphisms (SNPs) in glutamate-related genes have been associated with antipsychotic response or treatment resistance. A GWAS conducted by Stevenson *et al* [48] identified two SNPs in the *GRID2* gene (rs9307122 and rs1875705) that were associated with reduced response to antipsychotic treatment according to the Brief Psychiatric Rating Scale change score. The results found by Stevenson *et al* [48] support the hypothesis that genetic variation in glutamate system genes may impact the clinical trajectory of the patients treated with antipsychotic medications, and that these may represent a broader involvement of neurodevelopmental pathways. Furthermore, the *GRID2* gene is selectively expressed in Purkinje cells in the cerebellum where they play a key role in synaptogenesis, synaptic plasticity and motor coordination. For that matter, different mutations in *GRID2* have been shown to cause cerebellar ataxia in human [49]. In a study conducted by Schwenkreis *et al* [50] proved the existence of abnormal motor cortex activation by TMS in some types of genetically defined spinocerebellar ataxia (SCA), whereas other genetic subgroups show normal responses.

Adhesion G protein-coupled receptor B3 (*ADGRB3*) also known as *BAI3* is a gene that encodes a brain-specific angiogenesis inhibitor and is thought to be a member of the secretin receptor family. This gene play a key role in the regulation of several aspects of the central nervous system, such as axon guidance, myelination and synapse formation and function [51]. *ADGRB3* SNPs has already been associated with schizophrenia, bipolar disorder and drug addiction. In a family-based study conducted by Scuderi *et* al [38] shows a correlation between a disrupting intragenic duplication involving several exons of the *ADGRB3* and intellectual disability, cerebellar atrophy and behavioral disorder. The BAI proteins are highly expressed in the brain and have been identified at postsynaptic densities in the forebrain and cerebellum. The involvement of these proteins in the development of functional neuronal networks is related to their structural characteristics. The morphology and complexity of dendritic arborization allow functional differences of neurons, and deficits in neuronal morphogenesis correlate with psychiatric disorders. Lanoue *et al* [52] presented evidence both *in vivo* and in vitro for a signaling pathway regulating the morphogenesis of dendrites involving *BAI3*. The authors suggest that an interaction between BAI3 and the ELMO1 protein (important regulator of RAC1 RhoGTPase) is involved in this signaling.

*COL9A3* is a gene that encodes one of the three alpha chains of the type IX collagen. Mutations in this gene are associated with multiple epiphyseal dysplasia type 3. Some of the brain collagen proteins are expressed by neurons, suggesting their involvement in growth regulation and axonal orientation, synaptogenesis, cell adhesion, and brain architecture development [53]. Collagen biosynthesis in the brain can be abnormal in many hereditary diseases. Much of the brain pathology associated with collagen are related to neurodevelopment. Collagen type IV is known to inhibit glial differentiation in cortical cell cultures and to be enhanced in the frontal and temporal cortex of patients with Alzheimer’s disease [54,55].

Lymphocyte antigen 9 (*LY9*) belong to signaling lymphocytic activation molecule (SLAM) family of immunomodulatory receptors. According to previous studies, the activation of upstream gene regulatory pathways that modulate gene expression in immune cells may be linked to MDD [56]. Between the active pathways there are a family of transcription factors (TFs), the glucocorticoid receptor (GR), cAMP response element-binding (CREB), early growth response (EGR) family TFs, and pro-inflammatory cytokines, such as tumor necrosis factor-α (TNF-α), interleukin (IL)-1β and IL-6 [57–60]. In a study conducted by Mellon *et* al [61] to test the theory on transcriptional control pathways that may be active in MDD, the authors found that among the main negatively regulated transcripts in MDD patients are the cell surface antigens of leukocytes (CD6, CD7, CD22 and LY9). Differential expression of these transcripts may be associated with possible changes in the distribution of leukocyte subset in MDD patients.

*FOXN3* is a protein coding gene member of the forkhead/winged helix transcription factor family. Recent GWASs was conducted to analyze possible gene that are associated with suicide found significant correlation between evidence for suicidality and the gene *FOXN3* [62,63]. However, the way this gene may influence the risk of suicide is not fully elucidated.

## Conclusions

In this study, we set out to test whether polymorphic profiles are associated to rTMS treatment outcome. From the findings, we may consider that the responsiveness to rTMS may be associated to several pathways and not just to the influence of a single gene. As already reported in the literature the influence of genes such as *APP, GRID2, SPPL2A* and others on MDD (also described here), suggests that the genes found may influence the response to rTMS. However, the molecular mechanisms by which these genes may influence the response to rTMS treatment are unknown, requiring further investigation.

This study has some limitations that should be noted. The sample size used in this study was smaller than typically employed in genetic association studies and stratified (Australian patients with a diagnosis of major depression disorder refractory to pharmacological treatment). Although traditional GWAS require a vast number of genotyped individuals, this method is expensive and time-consuming. A potential solution for this is extreme phenotypic sampling. Recent studies have compared the results of extreme phenotypic sampling with large-scale samples, and showed that extreme phenotypes are effective [64]. This method allow to identify rare causal SNPs with increased efficiency. Due to heterogeneity of MDD the study with homogenous patient subgroups allows a better understanding about etiological mechanisms and thus the development of patient-specific treatment [65]. In addition, few reports have been found in the literature associating genetic profile and response to rTMS therapy in treatment-resistant depression patients [66–68]. Further replication is necessary to confirm the present findings and to further uncover the genetic profile of refractory individuals with MDD and their responsiveness to rTMS.

## Data Availability

Our ethics approval does not allow data sharing.

